# Immunotherapies for risk reduction in age-associated neurodegenerative diseases: impact of sex and treatment duration

**DOI:** 10.64898/2026.03.06.26347446

**Authors:** Helena Cortes-Flores, Georgina Torrandell-Haro, Roberta Diaz Brinton

**Affiliations:** Center for Innovation in Brain Science, University of Arizona, Tucson, AZ, United States; Department of Pharmacology, University of Arizona College of Medicine, Tucson, AZ, United States; Department of Neurology, University of Arizona College of Medicine, Tucson, AZ, United States

**Keywords:** Alzheimer’s Disease, Neurodegenerative Disease, anti-inflammatory treatment, NSAIDs, retrospective analysis

## Abstract

**Introduction:** Neurodegenerative diseases (NDDs) including Alzheimer’s disease (AD), Parkinson’s disease (PD), multiple sclerosis (MS), amyotrophic lateral sclerosis (ALS), and non-AD dementias share chronic neuroinflammatory mechanisms that contribute to neuronal injury and disease progression. While anti-inflammatory therapies (AITs) are associated with reduced neurodegenerative disease risk, knowledge regarding the impact of biological sex and treatment duration across multiple NDDs remains limited.

**Methods:** We conducted a retrospective cohort analysis using a large propensity-score-matched population (n = 190,308; 95,154 treated vs 95,154 untreated) to evaluate associations between long-term AIT exposure and incidence of major NDDs. Disease-specific and combined outcomes were assessed across drug classes (NSAIDs, corticosteroids, immunomodulators), sex, age, and therapy duration.

**Results:** AIT exposure was associated with a significantly lower risk of developing any NDD (RR = 0.47, 95% CI 0.43–0.48, p < .0001) and was equally effective in both sexes. Risk reduction was observed for each individual disease: AD (RR = 0.40), non-AD dementia (RR = 0.51), PD (RR = 0.43), MS (RR = 0.25), and ALS (RR = 0.48). Among drug classes, immunomodulators conferred the largest reduction (RR = 0.19), followed by corticosteroids (RR = 0.41) and NSAIDs (RR = 0.42). Duration analyses revealed a graded benefit, with RR declining from 0.94 (<1 year) to 0.25 (>6 years). Risk reduction was strongest in older participants (75–79 years).

**Discussion:** Chronic use of anti-inflammatory or immunomodulatory therapies was associated with substantially reduced incidence of multiple neurodegenerative diseases in both sexes. The strongest effects were observed with immunomodulator use and prolonged therapy duration, suggesting that sustained modulation of systemic inflammation confers broad neuroprotective effects in both sexes. These findings highlight the potential of targeting immune-inflammatory pathways for neurodegenerative disease prevention and can inform prospective mechanistic and interventional studies.

## 1 INTRODUCTION

Neurodegenerative diseases (NDDs) represent a significant and growing public health concern, with an estimated 100 million Americans affected and associated costs exceeding $800 billion annually in the United States(Gooch et al., 2017, 2024). As the aging population continues to expand, the incidence and burden of these diseases are projected to increase(2024). Among the most prevalent NDDs are Alzheimer’s disease (AD), Parkinson’s disease (PD), multiple sclerosis (MS), and amyotrophic lateral sclerosis (ALS).

Despite distinct etiologies and clinical profiles, these four disorders share fundamental pathophysiological mechanisms, including protein aggregation, mitochondrial dysfunction, oxidative stress, and chronic neuroinflammation (Gonzales et al., 2022, Lamptey et al., 2022). Accumulating evidence indicates that immune dysregulation and sustained activation of glial cells contribute to both the initiation and progression of NDDs(Gonzales et al., 2022, Lamptey et al., 2022, Heneka et al., 2015, Leng and Edison, 2021, Stephenson et al., 2018).

Neuroinflammation is increasingly recognized as a key modulator in neurodegenerative diseases. Immune signals influence microglial priming, and shape the neuroimmune environment through cytokine signaling cascades (Erickson and Banks, 2018, Varatharaj and Galea, 2017). Long-term inflammatory conditions and elevated circulating cytokines have been associated with an increased risk of developing NDDs, whereas the use of anti-inflammatory or immunomodulatory agents has been proposed as a potential strategy to mitigate this risk (Antwi et al., 2025, Calsolaro and Edison, 2016).

Anti-inflammatory drugs include a wide range of agents that suppress inflammation and immune overactivity through distinct mechanisms(Fine, 2013 Nov, Hsiao CJ, 2010, Dinarello, 2010). For the purposes of this study, drugs were categorized into four different classes: nonsteroidal anti-inflammatory drugs (NSAIDs), corticosteroids, immunomodulatory small molecules (immunomodulators), and biologic drugs. NSAIDs reduce inflammation by inhibiting cyclooxygenase (COX) enzymes, reducing the production of pro-inflammatory prostaglandins. NSAIDs primarily target COX-1 and COX-2, thereby decreasing peripheral and, for those that cross the BBB, central prostaglandin-mediated inflammatory signaling. Many NSAIDs are small and sufficiently lipophilic to cross the blood–brain barrier (BBB), allowing some degree of central anti-inflammatory action. They are widely used to manage acute and chronic pain, fever, and inflammation, especially in musculoskeletal conditions (Owais Qureshi, 2023).

Corticosteroids exert broad immunosuppressive effects and anti-inflammatory effects by modulating gene transcription, reducing pro-inflammatory cytokine, and inhibiting immune cell activation (Sharman, 2023). They act through intracellular glucocorticoid receptors to downregulate inflammatory pathways such as NF-κB and AP-1, and they reduce the activity of multiple immune cell types including T cells, macrophages, and eosinophils. Corticosteroids readily penetrate the BBB, with agents such as dexamethasone achieving particularly high central nervous system (CNS) exposure (Liu et al., 2013). They are commonly prescribed in acute exacerbations of chronic inflammatory diseases (e.g., asthma, inflammatory bowel disease), and as a general immunosuppressant in severe systemic inflammation conditions(Liu et al., 2013).

Immunomodulatory small molecules are targeted therapies that alter immune cell proliferation, differentiation, or function, often by inhibiting key signaling enzymes or metabolic pathways. Common mechanisms include inhibition of JAK-STAT signaling, S1P receptor modulation, or blockade of intracellular kinases such as PDE4 or BTK. These molecular targets reduce cytokine production, T-cell activation, or pathogenic immune cell trafficking (Strzelec et al., 2023, Schwartz et al., 2017, Zheng et al., 2021) . Because they are of low molecular weight and often lipophilic, many immunomodulatory small molecules can cross the BBB to exert effects within the CNS. These drugs are typically used to treat autoimmune diseases such as rheumatoid arthritis or psoriasis (Strzelec et al., 2023, Schwartz et al., 2017, Zheng et al., 2021). Although immunomodulatory agents can act by stimulating or suppressing immune responses, drugs included in these analyses are exclusively anti-inflammatory and used to attenuate excessive immune activation.

Biologic drugs are therapeutic products derived from living organisms that are typically large, complex molecules (U.S. Food and Drug Administration (FDA), 2020, Walsh, 2018). For this study, we focused on biologic anti-inflammatory drugs, which specifically target components of the immune system involved in inflammation, including pro-inflammatory cytokines and immune cell surface receptors (Becher et al., 2017, Smolen et al., 2016). Common therapeutic targets include TNF-α, IL-6, IL-1β, IL-17, and immune checkpoint molecules involved in cell–cell communication. In contrast to the other classes, biologic drugs do not effectively cross the BBB due to their large molecular size and limited permeability. They are primarily used for reducing pathological inflammation in autoimmune and chronic inflammatory diseases such as rheumatoid arthritis, inflammatory bowel disease or psoriasis (Becher et al., 2017, Smolen et al., 2016).

Epidemiological studies have shown that individuals receiving chronic anti-inflammatory treatment, such as NSAIDs and corticosteroids, or disease-modifying therapies for autoimmune diseases like immunomodulators, may exhibit lower incidence rates of neurodegenerative outcomes, although results have been inconsistent across studies and populations (Ali et al., 2019, Gupta et al., 2015, Wang et al., 2015). Heterogeneity in disease mechanisms, drug classes, and treatment durations likely contributes to these discrepancies. Moreover, sex differences in immune response and disease susceptibility suggest that therapeutic effects may vary between men and women, yet few large-scale analyses have addressed these interactions (Klein and Flanagan, 2016).

To clarify the potential neuroprotective role of AIT across the neurodegenerative spectrum, we conducted a retrospective cohort analysis assessing the association between chronic AIT use and the risk of developing major NDDs, including AD, PD, ALS, MS, and non-AD dementias. We further examined whether these associations differed by drug class, sex, or treatment duration. By integrating these analyses, this study aims to identify consistent epidemiological signals of reduced neurodegenerative risk, providing translational insight into how systemic modulation of inflammation may influence long-term brain health.

## 2 METHODS

### 2.1 Data Source

The study utilized a retrospective analysis of insurance claim records from the Mariner dataset. The Mariner dataset contains insurance claims data from all territories of the United States, predominantly covering the Southeastern region (Branigan et al., 2020, Torrandell-Haro et al., 2020). It includes information on patient demographics, prescription records, patient diagnoses, and procedures, organized under Current Procedural Terminology, International Classification of Diseases, Ninth Revision (ICD-9), and International Statistical Classification of Diseases and Related Health Problems, Tenth Revision (ICD-10) codes. The utilized database contained records of 170 million patients with claims spanning from 2010 to April 2021.

PearlDiver software was used to access the healthcare data, enabling the interaction with individual commercial, state-based Medicaid, Medicare stand-alone prescription drug plan, group Medicare Advantage, and individual Medicare Advantage data.

This study adheres to the Strengthening the Reporting of Observational Studies in Epidemiology (STROBE) reporting guideline. Approval was obtained from the University of Arizona Institutional Review Board. Informed consent requirements were waived due to the use of deidentified data.

### 2.2 Study Design and Variables

The study investigated the impact of anti-inflammatory drugs on the risk of age associated NDDs. A subset of 3,182,960 patients was selected from the Mariner database for analysis. Patients under 60 years of age, with a history of neurosurgery or brain cancer, or with a diagnosis of NDDs prior to the index date were excluded. Patients were included if they were continuously enrolled in medical and pharmacy insurance for a minimum of 6 months before and 3 years after the index date. The index date was defined as the initial drug prescription record for the treatment group and at least a 6-month period following the first patient claim record for the control group. Two study groups were defined based on therapeutic intervention: the treatment group included patients with at least one charge for an anti-inflammatory drug, and the control group included patients without any anti-inflammatory drug charge. Medications considered in this study included oral, IV, and injected anti-inflammatories with FDA approval. Therapies were identified by Drug Codes (**Table S1**) using the generic names. The outcome was defined as the incidence of NDDs, based on ICD-9 and ICD-10 codes (**Table S2**), at least 1 year after the index date. This time period allowed for the onboarding to therapy and to detect only long-term effects of anti-inflammatory drugs. Age in the treatment group was determined based on the age of first exposure to anti-inflammatory drugs. Following the methodology used in previous studies (Branigan et al., 2020, Torrandell-Haro et al., 2020), an assessment of comorbidities known to be related with NDD development was conducted. This analysis facilitated the creation of a logistic regression-based propensity score matched cohort for treatment and control populations. Relative risk was calculated by comparing these groups to the control group and included assessment for sex differences. Cumulative hazard ratios were constructed using the propensity score matched population. The population was divided in four different age groups (60-64, 65-69, 70-74, and 75-79), and cumulative hazard ratios for all combined NDDs and AD were generated. Additionally, the effect of AIT duration on the risk of NDDs was evaluated through analysis for different durations of therapy: <1 year, 1 to 3 years, 3 to 6 years, and >6 years, as explained in Kim et al. (Kim et al., 2021).

### 2.3 Statistical Analysis

Statistical analyses were conducted between April 1st and July 3rd, 2025. Firstly, to ensure comparability between the control and treatment groups, propensity score matching was applied following the methodologies described in Branigan et al. (Branigan et al., 2020) and Torrandell-Haro et al.(Torrandell-Haro et al., 2020). Before the matching process, logistic regression was utilized to estimate the likelihood of each patient receiving anti-inflammatory drugs, considering potential confounding variables such as age, sex, region, Charlson Comorbidity Index (CCI), and comorbidity claim records. Subsequently, the control and treatment populations were matched using the statistically significant confounding variables identified in the regression model.

Unpaired two-tailed t-tests or χ2 tests were performed, as appropriate, to determine the statistical significance between the control and treatment populations in continuous and categorical variables. A two-sided P < 0.05 was considered statistically significant.

## 3 RESULTS

A subset of 3,182,960 patients was selected from the Mariner dataset. Inclusion, exclusion, and enrollment criteria were met by 1,046,074 patients, who were divided into the control and treatment groups according to their anti-inflammatory prescription charges (**Figure 1**). After applying the propensity score match function, the study group included 190,308 patients (mean [standard deviation (SD)] age, 67 [4.8] years) of which 95,154 had at least one prescription charge of an anti-inflammatory drug, and 95,154 were not prescribed with any anti-inflammatory drug. The average follow-up time was 7.9 [2.7] years for the control group and 7.85 [3.4] years for the treatment group.

**Figure 1:**
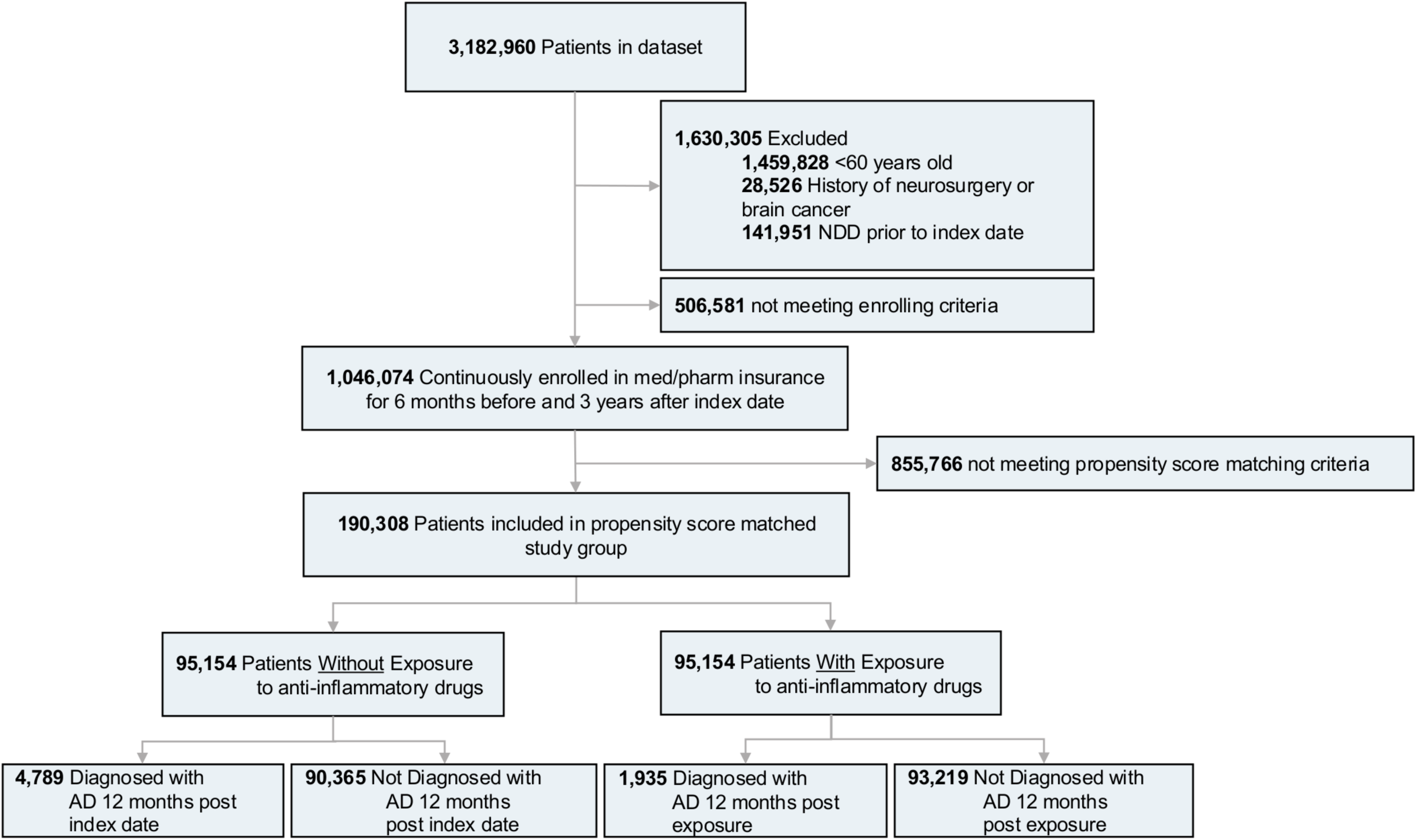
Study design and patient breakdown. NDD, neurodegenerative diseases; AD, Alzheimer’s disease.

In the unadjusted cohort there were statistically significant differences in demographic parameters, including sex and region of origin of patients. Similarly, comorbidity profiles between the control and treated groups were significantly different (**Table 1**). The treated cohort was younger than the control, with a higher percentage of the younger population (60 to 64 years old) (49.78% vs 20.42%) and a lower number of older patients (75 to 79 years old) (6.88% vs 21.40%). Additionally, the treated cohort was predominately female (56.62% vs 49.10%), while the control group had a higher percentage of males (50.90% vs 43.38%). The cohort receiving AIT exhibited a higher prevalence of all NDD-relevant comorbidities. However, CCI score indicated no significant differences between groups. In the propensity score matched population there were no significant differences in age, sex, region, and CCI when comparing the treated and control group, whereas significant differences in comorbidity diagnosis were conserved. Nevertheless, the percentage of patients in the treated group suffering from any NDD-relevant comorbidity decreased in comparison to the unadjusted cohort, with an average of 4.94% patients in the treated group having a diagnosis for any of the comorbidities assessed compared to 2.30% in the control group (**Table 1**).

**Table 1:**
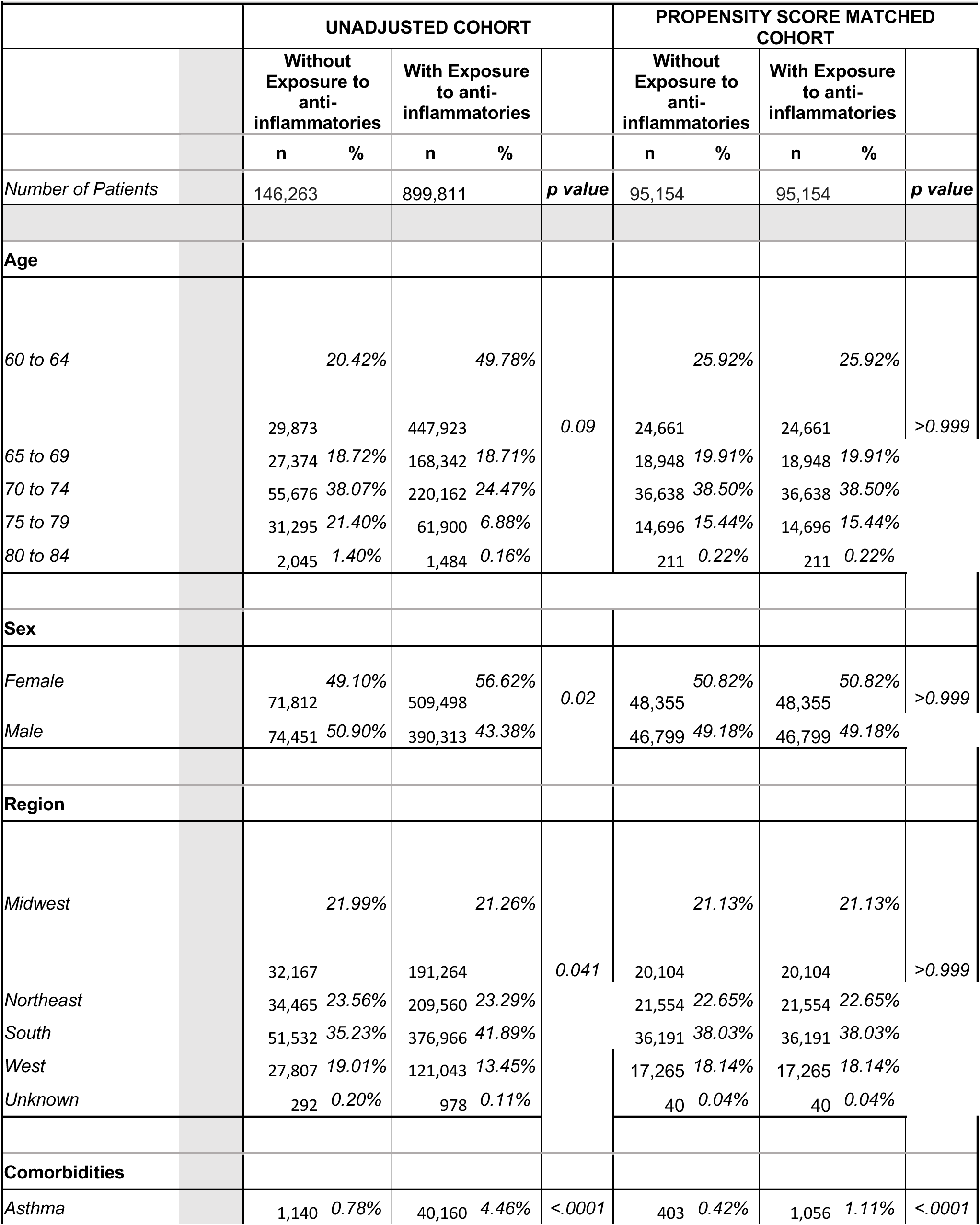
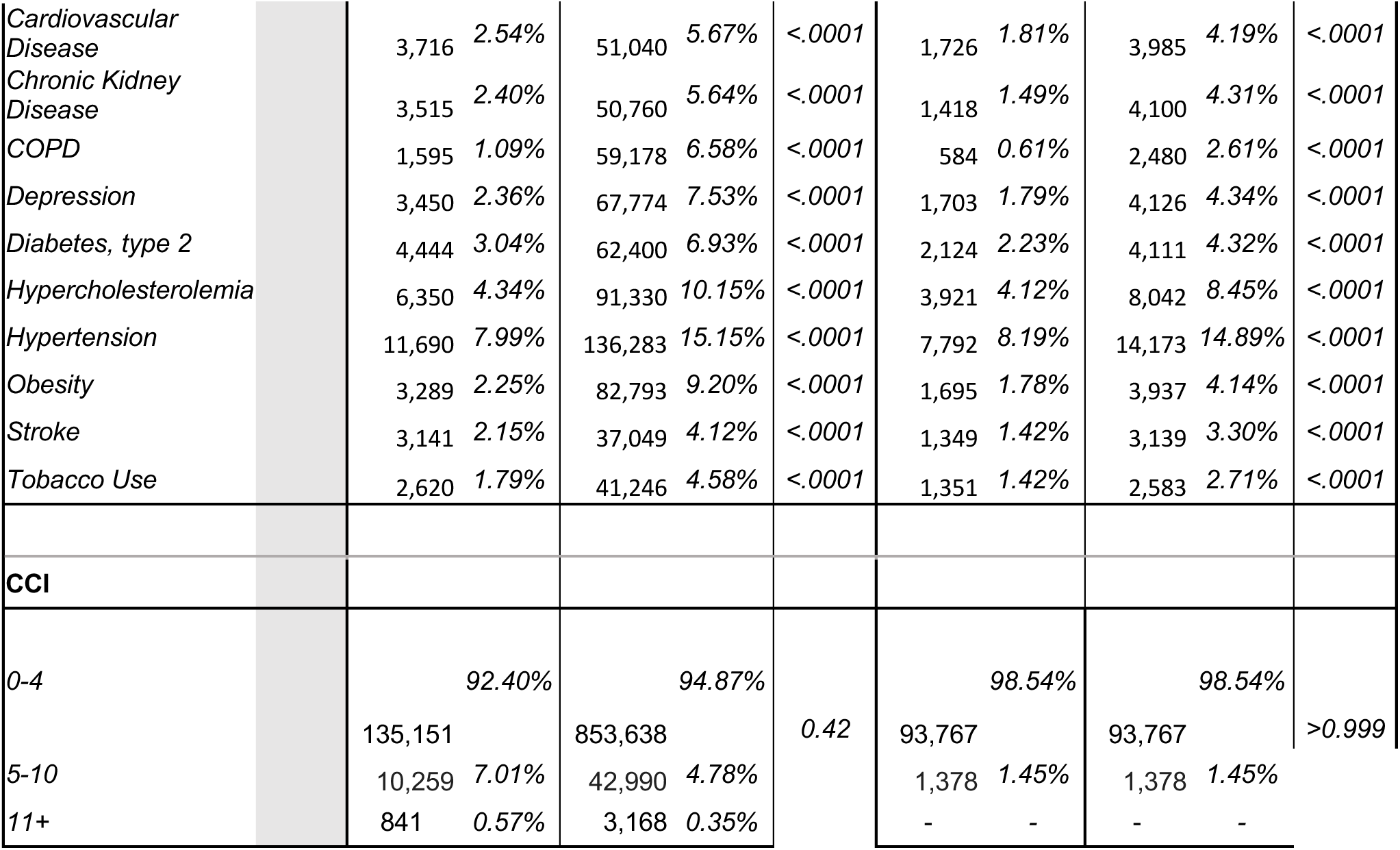
Baseline characteristics for unadjusted and propensity score-matched patients with or without exposure to anti-inflammatory drugs.

In the propensity score matched population, exposure to AIT was significantly associated with a decreased risk for all NDDs assessed when compared to the control group: AD (2.03 % vs 5.03 %, relative risk [RR]: 0.40; 95% confidence interval [CI]: 0.38-0.43; p < .0001), non-AD dementia (4.05 % vs 7.94 %, RR: 0.51; 95% CI: 0.49-0.53; p < .0001), PD (0.87% vs 2.04 %, RR: 0.43; 95% CI: 0.40-0.46; p < .0001), MS (0.03% vs 0.11 %, RR: 0.25; 95% CI: 0.17-0.39; p < .0001), ALS (0.05% vs 0.10%, RR: 0.48; 95% CI: 0.34-0.69; p < .0001), and combined NDDs (6.67 % vs 14.23%, RR: 0.47; 95% CI: 0.43-0.48; p < .0001) (**Table 2**) (**Figure 2A**). The number of patients needed to treat (NNT) to prevent one AD event was 33; 26 for non-AD dementia; 86 for PD; 1,204 for MS; 2,025 for ALS; and 13 for all NDDs combined (**Table 2**). Sex stratification indicated a slight significant difference only when analyzing all NDDs combined, with females receiving a marginally higher benefit in risk reduction (RR: 0.45; 95% CI: 0.44-0.46; p < .0001) compared to males (RR: 0.49; 95% CI: 0.47-0.52; p < .0001) (**Figure 2B**).

**Figure 2:**
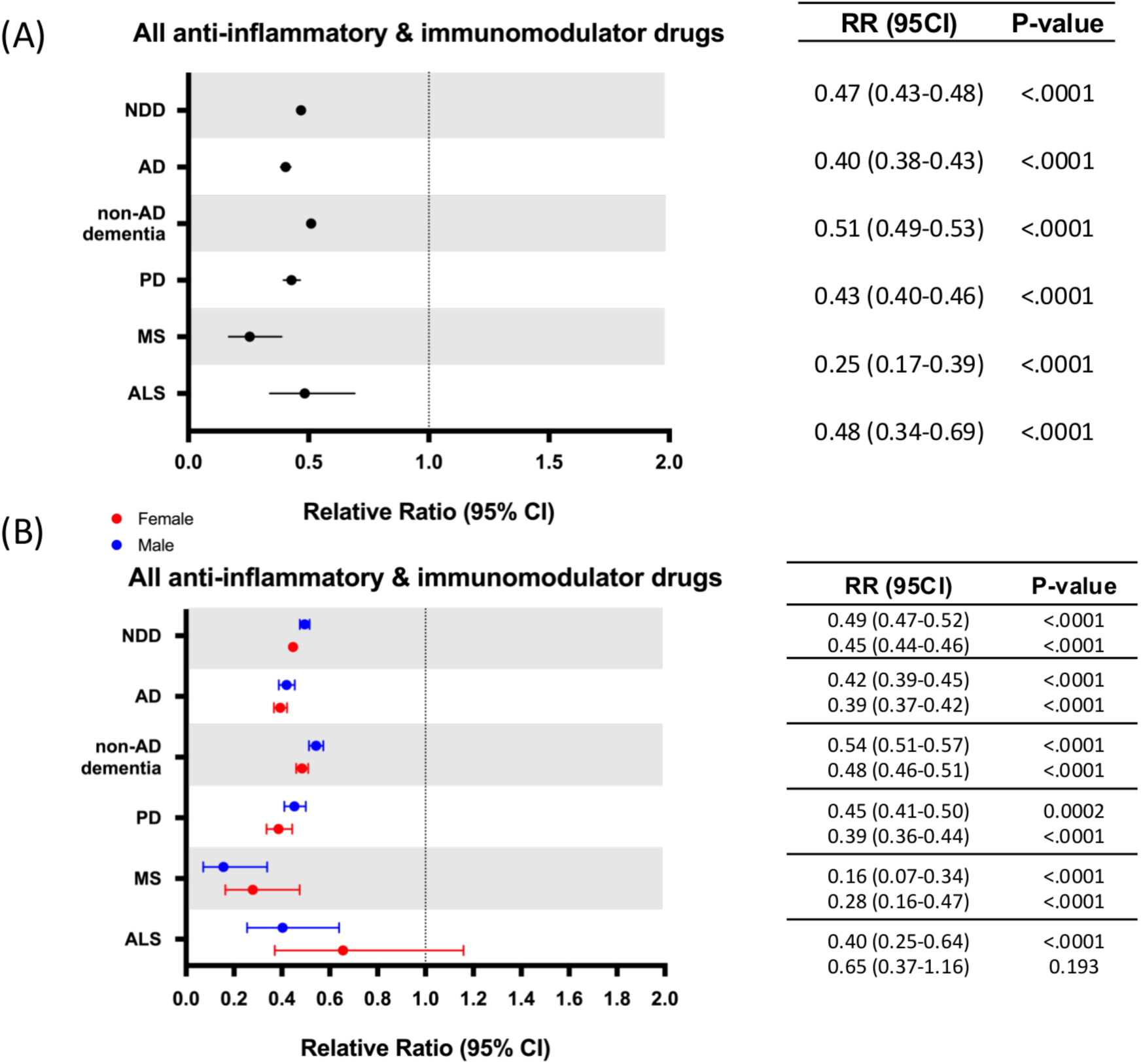
Relative risk of neurodegenerative diseases (NDDs) following anti-inflammatory drug use. (A) Relative risk of propensity score matched patients developing NDDs after receiving anti-inflammatory drugs. (B) Sex differences on relative risk of propensity score matched patients developing NDDs after receiving anti-inflammatory drugs. AD, Alzheimer’s disease; ALS, amyotrophic lateral sclerosis; CI, confidence interval; MS, multiple sclerosis; NDD, neurodegenerative diseases; PD, Parkinson’s disease; RR, relative risk.

**Table 2:**
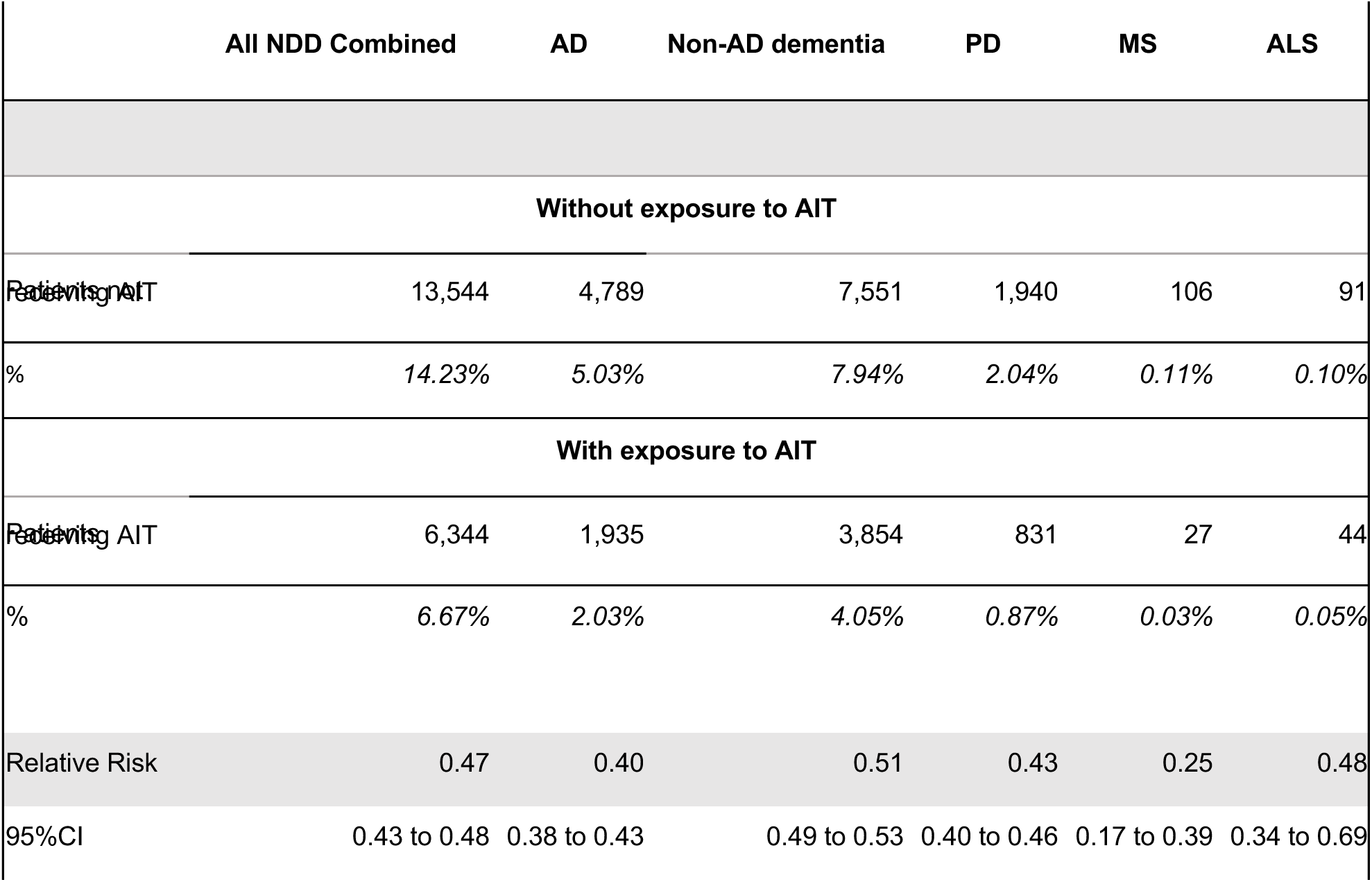
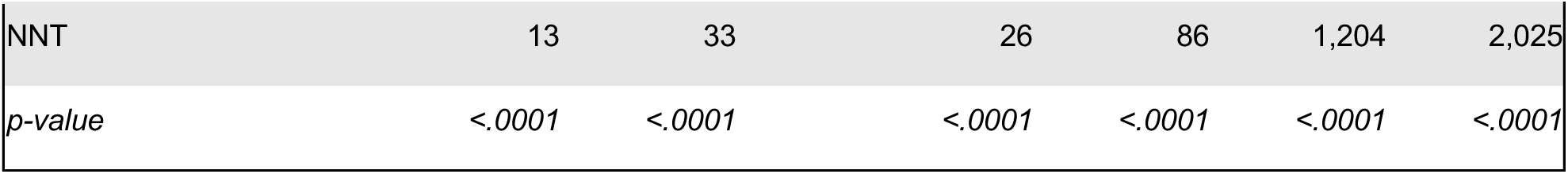
Relative risk of propensity score matched patients developing NDDs after receiving anti-inflammatory drugs. AD, Alzheimer’s disease; ALS, amyotrophic lateral sclerosis; CI, confidence interval; MS, multiple sclerosis; NDD, neurodegenerative diseases; NNT, number needed to treat; PD, Parkinson’s disease.

The treated population was divided into four groups according to the different drug classes included in the study: NSAIDs, corticosteroids, immunomodulators and biologic drugs. Most patients were prescribed corticosteroids (75,572 [79.42%]) and NSAIDs (71,469 [75.11%), followed by a smaller subset of patients prescribed with immunomodulators (3,214 [3.38%]). (**Figure 3A**). Given the small sample size of patients prescribed with biologic drugs (408 [0.43%]), this drug class was not included in analyses showed in figures 3 and 4.

**Figure 3:**
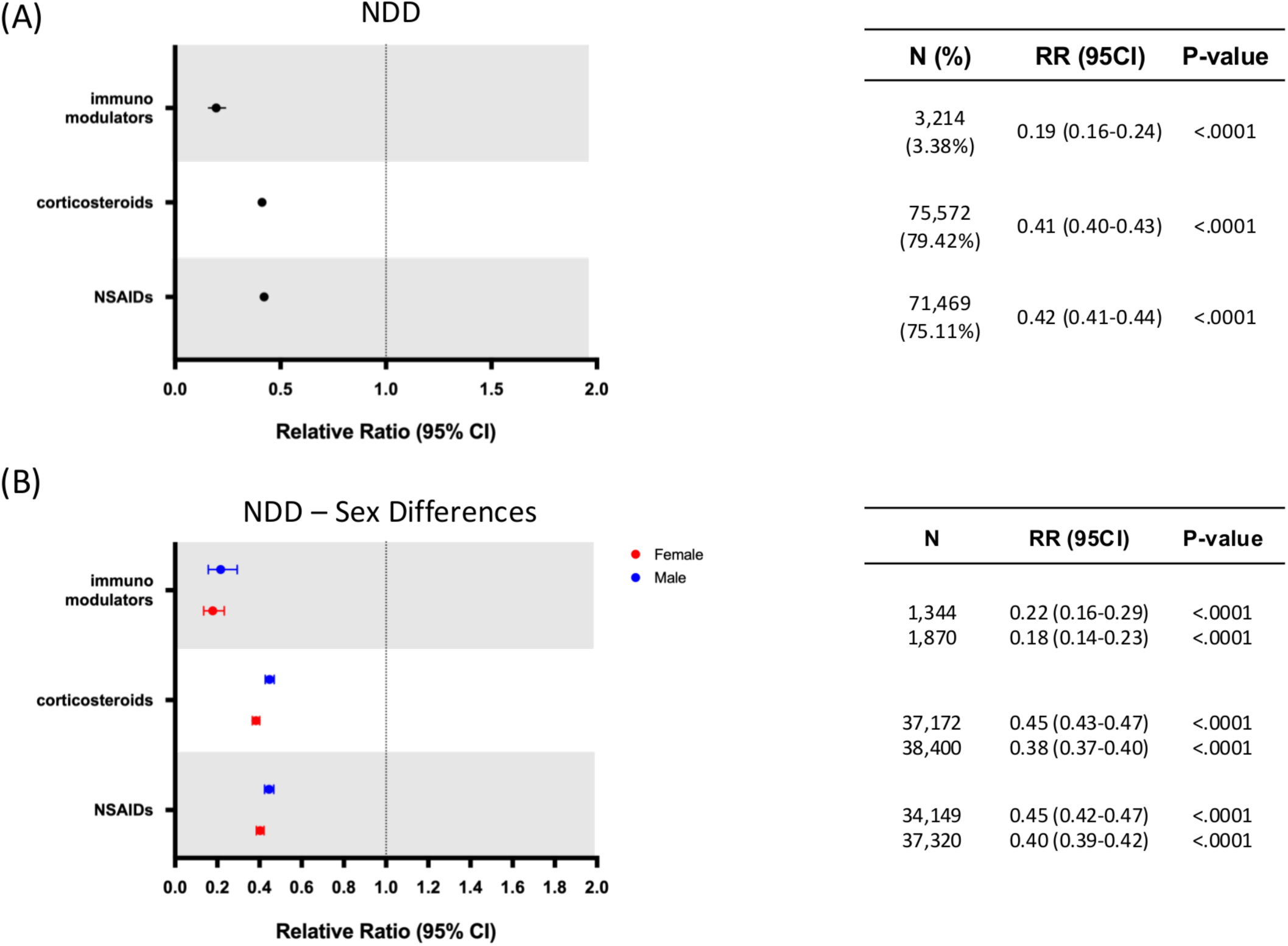
Relative risk of neurodegenerative diseases (NDDs) by anti-inflammatory drug class and sex. (A) Relative risk of developing NDD in patients with exposure to different classes of anti-inflammatory drugs. (B) Sex differences on relative risk of developing NDD in patients with exposure to different classes of anti-inflammatory drugs. CI, confidence interval; NDD, neurodegenerative disease; RR, relative risk. N = Number of patients receiving each drug class.

**Figure 4:**
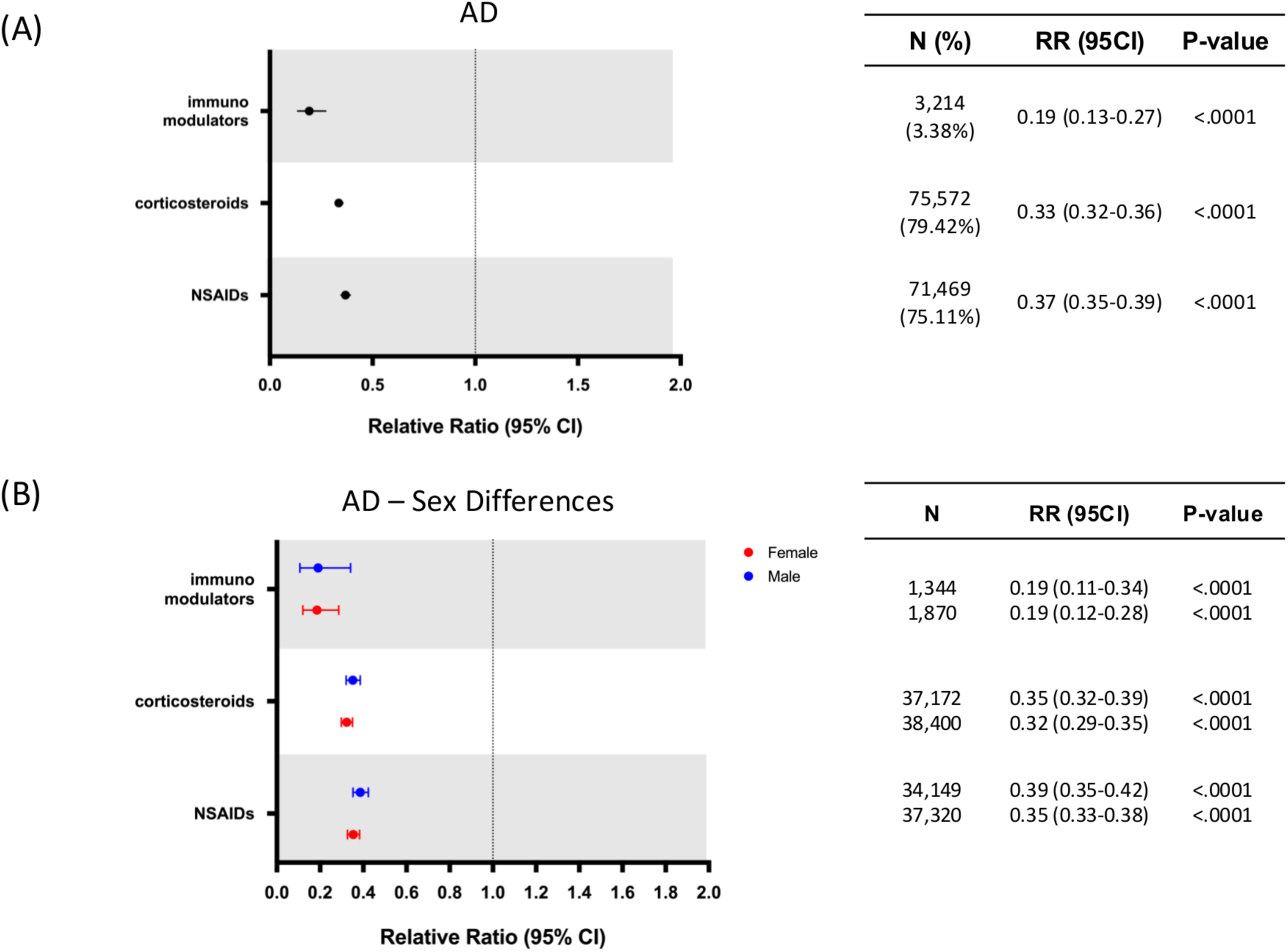
Relative risk of Alzheimer’s disease (AD) by anti-inflammatory drug class and sex. (A) Relative risk of developing AD in patients with exposure to different classes of anti-inflammatory drugs. (B) Sex differences on relative risk of developing AD in patients with exposure to different classes of anti-inflammatory drugs. AD, Alzheimer’s Disease; CI, confidence interval; RR, relative risk. N = Number of patients receiving each drug class.

Drug-class specific analyses for the NDD outcome (**Figure 3A**) revealed that all three major AIT classes (NSAIDs, corticosteroids, immunomodulators) were associated with risk reductions: immunomodulators exhibited the largest effect (RR: 0.19; 95% CI: 0.16-0.24; p < .0001), followed by corticosteroids (RR: 0.41; 95% CI: 0.40-0.43; p < .0001) and NSAIDs (RR: 0.42; 95% CI: 0.41-0.44; p < .0001) (all p < .0001). Sex-stratified analyses (**Figure 3B**) indicated consistent benefit in both females and males for immunomodulator and NSAID drug classes, whereas for corticosteroids females exhibited a slightly greater benefit in NDD risk reduction (RR: 0.38; 95% CI: 0.37-0.40; p < .0001) compared to males (RR = 0.45; 95% CI: 0.43-0.47; p < .0001).

Specific drug-class analysis for AD exhibited similar results with immunomodulators exhibiting the highest risk reduction (RR: 0.19; 95% CI: 0.13-0.27; p < .0001), followed by corticosteroids (RR: 0.33; 95% CI: 0.32-0.36; p < .0001), and NSAIDs (RR: 0.37; 95% CI: 0.35-0.39; p < .0001) (**Figure 4A**). Sex stratification for AD risk reduction for each treated group indicated that both females and males exhibited comparable risk reduction profiles (**Figure 4B**).

Age emerged as a risk modifier for all NDDs combined and AD risk specifically, as depicted in the cumulative hazard ratio curves (**Figure 5**). Cumulative hazard ratio curves were computed using the propensity score matched population to assess disease development over time. At a younger age (60 to 64 years old), when age-related NDDs are not likely to emerge, exposure to AIT did not exhibit an impact on NDD and AD risk compared to the control group. In contrast, a progressively greater distinction appeared between the two groups with advancing age, where the non-exposure group exhibited an elevated risk for NDD and AD development compared to the treatment group. The oldest age range of patients assessed in the study (75 to 79 years old), which corresponds with the demographic at the highest risk of developing AD, exhibited the most pronounced divergence in the cumulative hazard plots (**Figure 5**). This outcome indicated that patients aged 75 to 79 years of age exhibited the greatest benefit of AIT in reducing risk for NDDs, and AD specifically. Additionally, sex differences emerged in older patients (> 70 years) in the control group, with non-treated females exhibiting a higher rate of disease conversion for NDDs and AD compared to males (**Figure 6**). Interestingly, sex differences were not detected in the treated population, where females and males displayed comparable risk of NDD and AD development over time (**Figure 6**).

**Figure 5:**
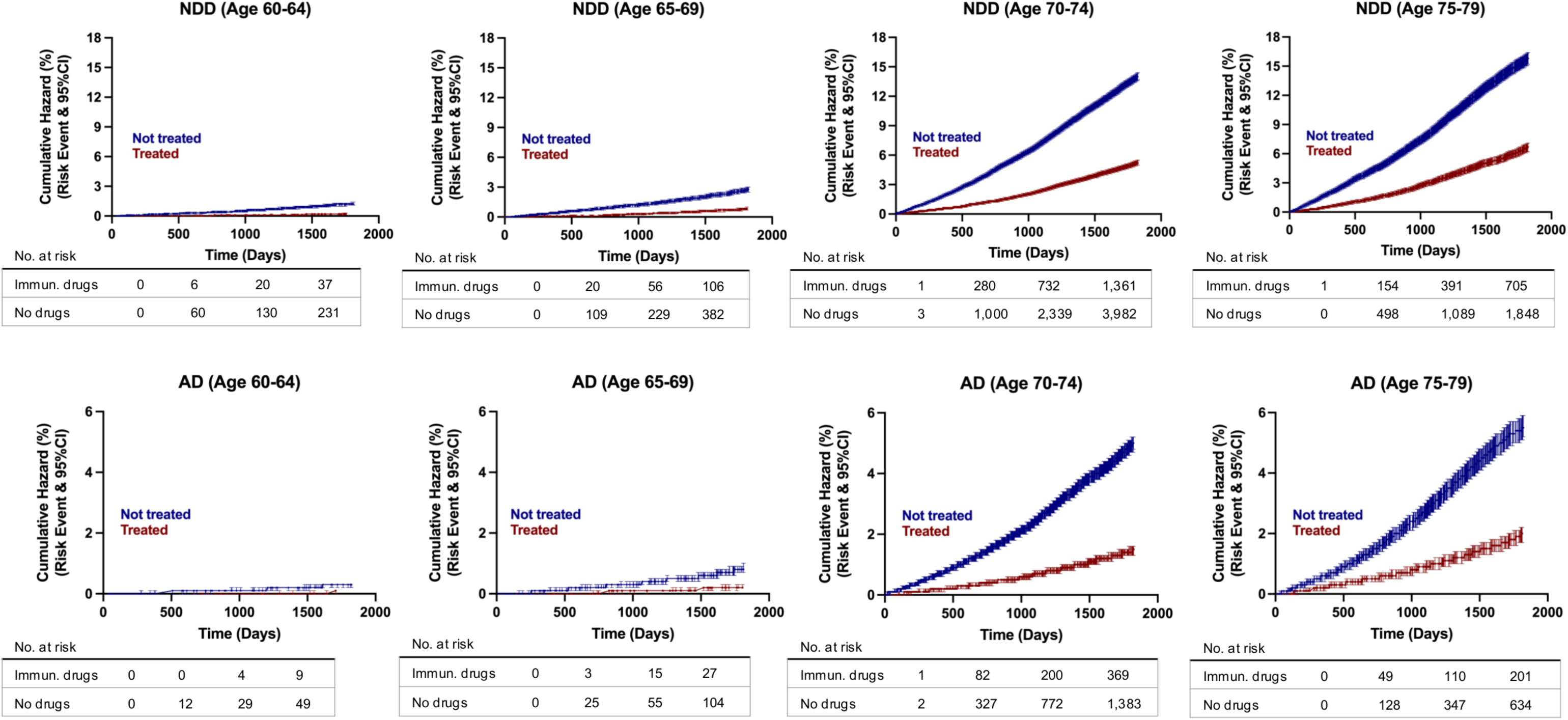
Hazard ratio curves for risk of developing AD and NDDs combined in propensity score matched patients. AD, Alzheimer’s disease; NDD, neurodegenerative disease.

**Figure 6:**
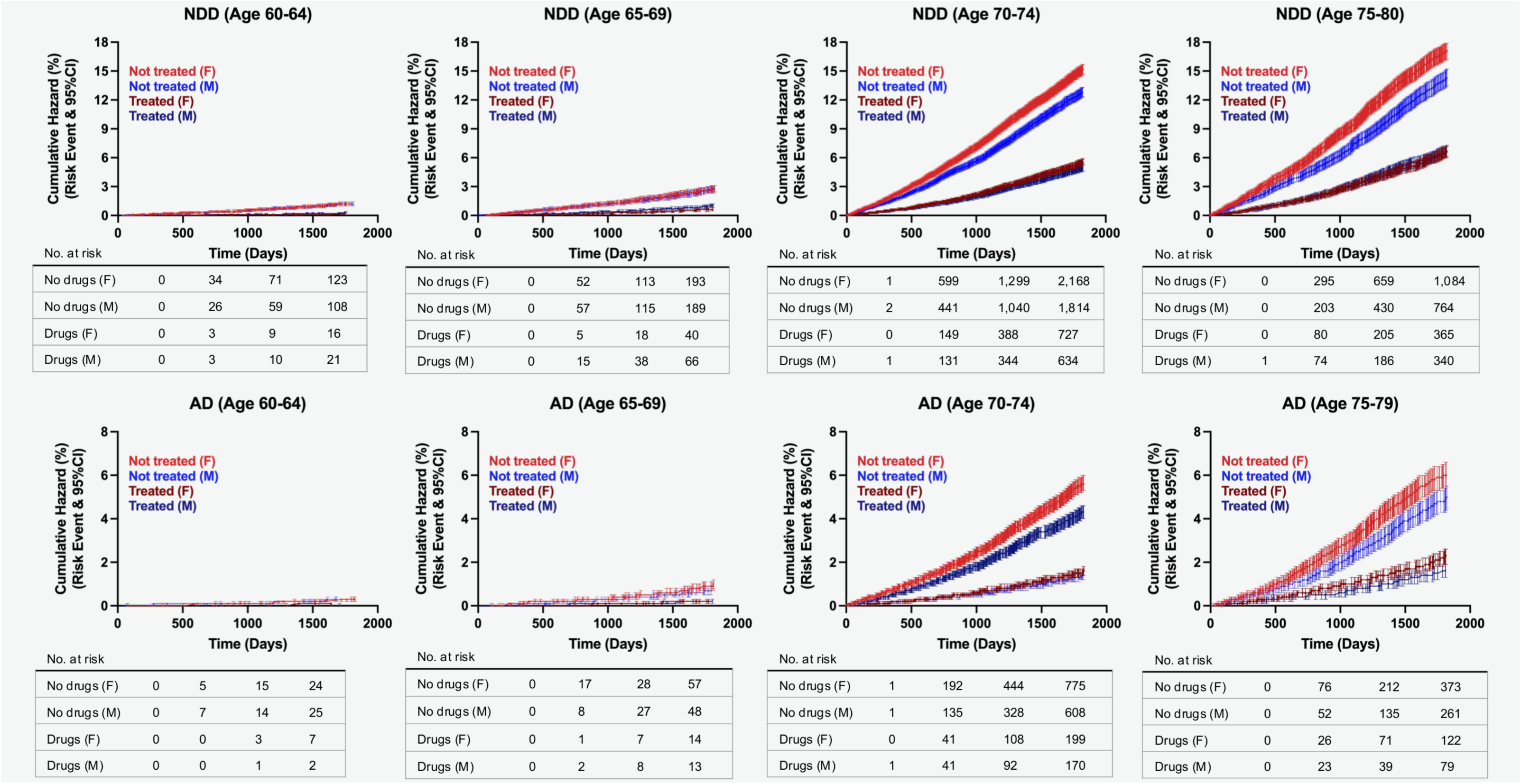
Hazard ratio curves for risk of developing AD and NDDs combined in propensity score matched patients, females versus males. AD, Alzheimer’s disease; NDD, neurodegenerative disease.

The impact of duration of AIT was assessed in the propensity score matched group for NDDs and AD. Within the treated population most patients were prescribed with an AIT for longer than 6 years: 4,128 (4.34%) patients received AIT for 1 year or less, 12,436 (13.07%) for 1 to 3 years, 29,346 (30.84%) for 3 to 6 years, and 47,199 (49.60%) for 6 years or longer. Duration of therapy analyses for the combined NDD outcome indicated a progressive reduction in RR with increasing treatment duration, with statistically significant reductions of risk only for treatments exceeding 1 year: <1 year (RR: 0.94; 95% CI: 0.87-1.02; p = 0.945), 1-3 years (RR: 0.92; 95% CI: 0.88-0.97; p = 0.0007), 3-6 years (RR: 0.58; 95% CI: 0.56-0.60; p < 0.0001), >6 years (RR: 0.25; 95% CI: 0.24-0.27; p < 0.0001) (**Figure 7A**). Sex-stratified duration analysis indicated comparable effect sizes in females and males, with a slightly greater benefit in females for treatment >6 years (Males: RR: 0.28; 95% CI: 0.26-0.30; p < 0.0001 vs Females: RR: 0.23; 95% CI: 0.22-0.25; p < 0.0001) (**Figure 7B**).

**Figure 7:**
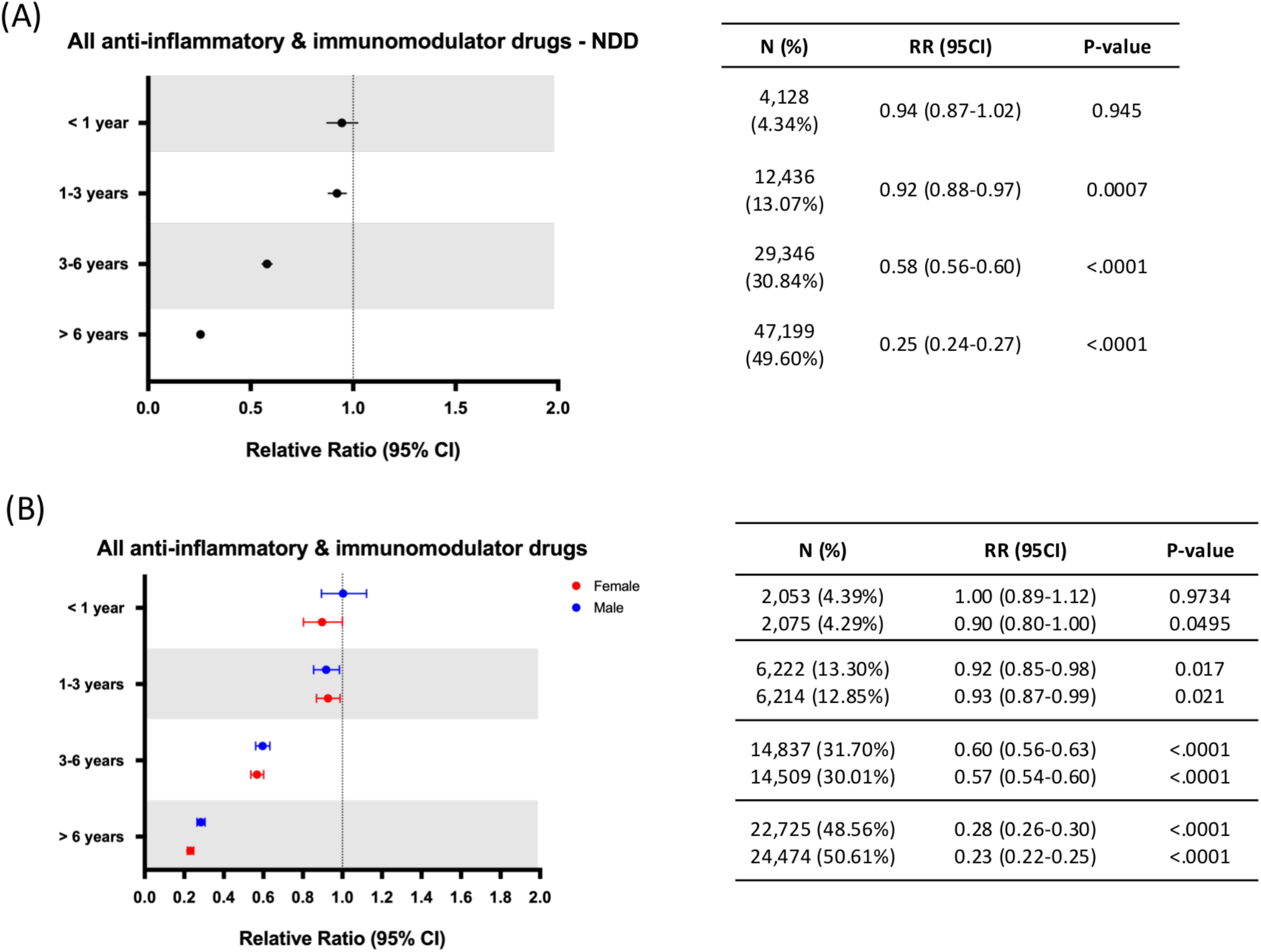
Relative risk of neurodegenerative diseases (NDDs) by anti-inflammatory therapy duration. CI, confidence interval; RR: relative risk; NDD, neurodegenerative disease; N = Number of patients receiving AIT for each of the time periods assessed.

Focused analysis on AD exhibited a similar profile where treatment for 1 year or less did not significantly modify the relative risk for development of AD (RR: 0.94; 95% CI: 0.82-1.08; p = 0.402), while all treatment durations greater than 1 year were associated with significant AD risk reduction. Additionally, longer AIT durations were associated with greater AD risk reduction, where treatment for 6 years and longer was associated with maximal AD risk reduction (RR: 0.21; 95% CI: 0.19-0.23; p < .0001), followed by treatment for 3 to 6 years (RR: 0.50; 95% CI: 0.46-0.53; p < .0001) and treatment for 1 to 3 years (RR: 0.82; 95% CI: 0.75-0.90; p < .0001) (**Figure 8A**). Sex differences were also comparable to NDD results, with no differences emerging for any treatment duration except for a slightly higher risk reduction in females observed only for treatments of 6 years or longer (Males: RR: 0.24; 95% CI: 0.21-0.28; p = 0.0002 vs Females: RR: 0.18; 95% CI: 0.16-0.20; p < 0.0001) (**Figure 8B**).

**Figure 8:**
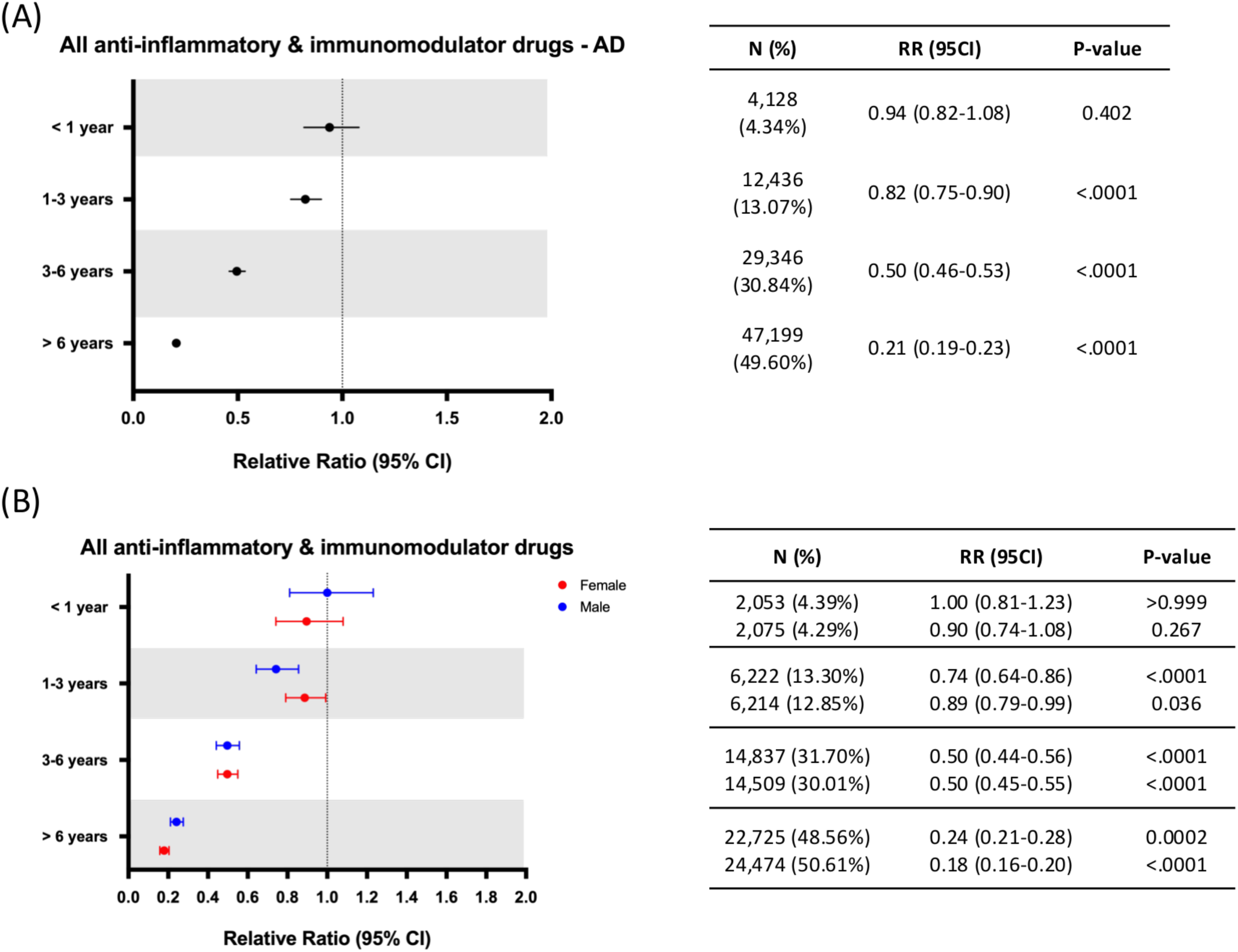
Relative risk of Alzheimer’s disease (AD) by anti-inflammatory therapy duration. AD, Alzheimer’s disease; CI, confidence interval; RR: relative risk. N = Number of patients receiving AIT for each of the time periods assessed.

## 4 DISCUSSION

This retrospective cohort study provided evidence that AIT is associated with substantially reduced incidence not only of AD but also a broader spectrum of NDDs including non-AD dementia, PD, MS and ALS. Importantly, the magnitude of association appears greater with longer duration of therapy and within the older age brackets, and holds across sexes and drug classes (though with variation). These findings extend prior observational work focused largely on AD and suggest that modulation of systemic and central immune/inflammatory processes may confer broad neuroprotective benefits. As inflammation is the unifying mechanism of NDDs, targeting inflammatory processes represents a common therapeutic strategy across these disorders, further supporting the biological plausibility of these associations.

The concept of neuroinflammation as a contributor to NDD pathogenesis has become increasingly relevant over the last decade. Chronic activation of microglia and astrocytes, and the release of pro-inflammatory cytokines (e.g., TNF-α, IL-1β, IL-6) have been implicated in AD, PD, ALS and other disorders (Kwon and Koh, 2020, Zhang et al., 2023). For example, glial activation and immune system dysregulation have been found in human post-mortem studies and animal models across all major NDDs (Chen et al., 2016, Zhang et al., 2023). The fact that numerous NDDs share immune/inflammatory components supports the concept of a shared mechanism of NDD, and thus shared therapeutic target, specifically anti-inflammatory interventions.

The study population was propensity score matched to reduce potential biases between patients exposed to anti-inflammatory drugs and those with no exposure. The two cohorts presented no differences in age, sex, and region, whereas differences in their comorbidity profile remained, with the control group being healthier than the treated group. Notably, despite the untreated group having a lower overall comorbidity burden, the incidence of NDDs was higher in this cohort compared to the treated group, suggesting a robust protective effect of AIT in reducing NDD risk. CCI values exhibited no significant differences between the control and treated group, and most patients (98.54%) fell under a CCI score of 0-4, which corresponds to the healthiest status.

Data indicating that immunomodulators exhibited the greatest risk reduction compared to corticosteroids and NSAIDs aligns with mechanistic pathways as immunomodulators are often prescribed for systemic autoimmune/inflammatory conditions and may induce greater and sustained immune modulation, including regulation of cytokine signaling, lymphocyte activation, and microglial priming, compared to classical anti-inflammatory agents (Faquetti et al., 2024, Wieczorek and Strosznajder, 2024, Owais Qureshi, 2023). Preclinical studies indicate that targeting immune pathways (e.g., JAK/STAT, sphingosine-1-phosphate receptors) can impact neurodegeneration in models of AD and MS (Ashraf et al., 2022, Palumbo et al., 2021). Although sample size for immunomodulator exposure was smaller, the effect size and consistency across sexes support its potential relevance.

The clear pattern of increasing benefit with greater duration of treatment supports the hypothesis that sustained immune–inflammatory modulation is required to impact long-latency neurodegenerative processes. This mirrors previous work indicating that short-term AIT (e.g., <15 months) did not reduce AD risk in randomized trials, whereas long-term observational use was beneficial (Ali et al., 2019, Gupta et al., 2015, Wang et al., 2015, Kwon and Koh, 2020, Zhang et al., 2023) . The greater effects in older age groups (75-79 yrs) may reflect a higher baseline risk of NDDs and therefore a greater absolute risk reduction induced by AIT intervention. At younger ages (60-64 yrs), the low incidence of NDD likely reduces an observable effect. Alternatively, it may reflect a “window of opportunity” where long-term AIT initiated later still provides benefit, perhaps by dampening accumulating inflammatory burden.

The absence of strong sex differences in relative risk across drug classes, durations and age is notable given established sex differences in immune function (females generally exhibit stronger immune responses) and NDD incidence (Klein and Flanagan, 2016). Our data suggest that AIT benefit is comparable in both sexes. However, the higher conversion rates in untreated females compared to males was most significant in the older age groups. The difference in conversion rates was mitigated by AIT. This may indicate that inflammation constitutes a more prominent modifiable risk in females, but when treated, the effect balances across sexes.

The finding of risk reduction not only for AD but also for PD, MS and ALS indicates that immune/inflammatory modulation has an impact upstream of and across multiple pathogenic pathways (protein aggregation, synaptic loss, motor neuron vulnerability) rather than disease-specific mechanisms alone. This aligns with the growing view that systemic inflammation and microglial/astroglial activation represent a common feature of neurodegeneration (Castro-Gomez and Heneka, 2024, Giri et al., 2024).

From a translational perspective, these results raise the possibility that long-term use of anti-inflammatory / immunomodulatory therapies may serve as a preventive strategy for neurodegenerative disease. While RCTs to date have not shown therapeutic efficacy for NDD treatment (Miguel-Alvarez et al., 2015), findings reported herein provide a broad timeframe for prevention and age frame for maximal preventive impact.

Outcomes of these analyses suggest that targeted prevention trials in persons at high risk for age-associated neurodegenerative diseases could benefit from immunotherapy, particularly immunomodulators, introduced early in the aging process and maintained over longer treatment durations.

### Limitations

Several limitations apply to this study. First, as with all retrospective cohort analyses, causality cannot be established, and additional parameters that could not be accounted for (e.g., AIT indication, socioeconomic status, over-the-counter AIT) may bias results. Second, the claims dataset lacks detailed clinical characterization (e.g., laboratory biomarkers, disease subtype, severity) and may misclassify diagnoses (particularly non-AD dementias). Third, the treated and control groups differed in comorbidity profiles prior to matching (although CCI was balanced), and patients receiving AIT likely represent individuals with chronic inflammatory or autoimmune conditions that may themselves influence NDD risk (either increasing or decreasing). Fourth, the sample size for some diseases (especially MS, ALS) and for immunomodulator subgroup is small. Finally, we cannot account for potential off-label or over-the-counter AIT use in control group.

## Author Contribution

HC-F: Conceptualization, Formal analysis, Writing – original draft, Writing – review & editing. GT-H: Conceptualization, Supervision, Writing – review & editing. RB: Conceptualization, Funding acquisition, Resources, Supervision, Writing – review & editing.

## Data Availability Statement

The datasets presented in this article are not readily available because restrictions apply to the availability of some or all data generated or analyzed during this study to preserve patient confidentiality or because they were used under license. The corresponding author will on request detail the restrictions and any conditions under which access to some data may be provided. Requests to access the datasets should be directed to.

## Reference

2024. 2024 Alzheimer’s disease facts and figures. Alzheimers Dement, 20, 3708–3821.

Ali, M. M., Ghouri, R. G., Ans, A. H., Akbar, A. & Toheed, A. 2019. Recommendations for Anti-inflammatory Treatments in Alzheimer’s Disease: A Comprehensive Review of the Literature. Cureus, 11, e4620.

Antwi, M. H., Bockarie, A., Osei, G. N., Donkor, D. M. & Simpong, D. L. 2025. Cytokines and immune biomarkers in neurodegeneration and cognitive function: A systematic review among individuals of African ancestry. Alzheimers Dement, 21, e70514.

Ashraf, H., Solla, P. & Sechi, L. A. 2022. Current Advancement of Immunomodulatory Drugs as Potential Pharmacotherapies for Autoimmunity Based Neurological Diseases. Pharmaceuticals (Basel), 15.

Becher, B., Spath, S. & Goverman, J. 2017. Cytokine networks in neuroinflammation. Nat Rev Immunol, 17, 49–59.

Branigan, G. L., Soto, M., Neumayer, L., Rodgers, K. & Brinton, R. D. 2020. Association Between Hormone-Modulating Breast Cancer Therapies and Incidence of Neurodegenerative Outcomes for Women With Breast Cancer. JAMA Netw Open, 3, e201541.

Calsolaro, V. & Edison, P. 2016. Neuroinflammation in Alzheimer’s disease: Current evidence and future directions. Alzheimers Dement, 12, 719–32.

Castro-Gomez, S. & Heneka, M. T. 2024. Innate immune activation in neurodegenerative diseases. Immunity, 57, 790–814.

Chen, W. W., Zhang, X. & Huang, W. J. 2016. Role of neuroinflammation in neurodegenerative diseases (Review). Mol Med Rep, 13, 3391–6.

Dinarello, C. A. 2010. Anti-inflammatory Agents: Present and Future. Cell, 140, 935–50.

Erickson, M. A. & Banks, W. A. 2018. Neuroimmune Axes of the Blood-Brain Barriers and Blood-Brain Interfaces: Bases for Physiological Regulation, Disease States, and Pharmacological Interventions. Pharmacol Rev, 70, 278–314.

Faquetti, M. L., Slappendel, L., Bigonne, H., Grisoni, F., Schneider, P., Aichinger, G., Schneider, G., Sturla, S. J. & Burden, A. M. 2024. Baricitinib and tofacitinib off-target profile, with a focus on Alzheimer’s disease. Alzheimers Dement (N Y), 10, e12445.

Fine, M. 2013 Nov. Quantifying the impact of NSAID-associated adverse events. . Am J Manag Care, s267–72.

Giri, P. M., Banerjee, A., Ghosal, A. & Layek, B. 2024. Neuroinflammation in Neurodegenerative Disorders: Current Knowledge and Therapeutic Implications. Int J Mol Sci, 25.

Gonzales, M. M., Garbarino, V. R., Pollet, E., Palavicini, J. P., Kellogg, D. L., JR., Kraig, E. & Orr, M. E. 2022. Biological aging processes underlying cognitive decline and neurodegenerative disease. J Clin Invest, 132.

Gooch, C. L., Pracht, E. & Borenstein, A. R. 2017. The burden of neurological disease in the United States: A summary report and call to action. Ann Neurol, 81, 479–484.

Gupta, P. P., Pandey, R. D., Jha, D., Shrivastav, V. & Kumar, S. 2015. Role of traditional nonsteroidal anti-inflammatory drugs in Alzheimer’s disease: a meta-analysis of randomized clinical trials. Am J Alzheimers Dis Other Demen, 30, 178–82.

Heneka, M. T., Carson, M. J., El Khoury, J., Landreth, G. E., Brosseron, F., Feinstein, D. L., Jacobs, A. H., Wyss-Coray, T., Vitorica, J., Ransohoff, R. M., Herrup, K., Frautschy, S. A., Finsen, B., Brown, G. C., Verkhratsky, A., Yamanaka, K., Koistinaho, J., Latz, E., Halle, A., Petzold, G. C., Town, T., Morgan, D., Shinohara, M. L., Perry, V. H., Holmes, C., Bazan, N. G., Brooks, D. J., Hunot, S., Joseph, B., Deigendesch, N., Garaschuk, O., Boddeke, E., Dinarello, C. A., Breitner, J. C., Cole, G. M., Golenbock, D. T. & Kummer, M. P. 2015. Neuroinflammation in Alzheimer’s disease. Lancet Neurol, 14, 388–405.

Hsiao CJ, C. D., Beatty PC, Rechtsteiner EA. 2010. National Ambulatory Medical Care Survey: 2007 summary. . Natl Health Stat Report., 1-32.

Kim, Y. J., Soto, M., Branigan, G. L., Rodgers, K. & Brinton, R. D. 2021. Association between menopausal hormone therapy and risk of neurodegenerative diseases: Implications for precision hormone therapy. Alzheimers Dement (N Y), 7, e12174.

Klein, S. L. & Flanagan, K. L. 2016. Sex differences in immune responses. Nat Rev Immunol, 16, 626–38.

Kwon, H. S. & Koh, S. H. 2020. Neuroinflammation in neurodegenerative disorders: the roles of microglia and astrocytes. Transl Neurodegener, 9, 42.

Lamptey, R. N. L., Chaulagain, B., Trivedi, R., Gothwal, A., Layek, B. & Singh, J. 2022. A Review of the Common Neurodegenerative Disorders: Current Therapeutic Approaches and the Potential Role of Nanotherapeutics. Int J Mol Sci, 23.

Leng, F. & Edison, P. 2021. Neuroinflammation and microglial activation in Alzheimer disease: where do we go from here? Nat Rev Neurol, 17, 157–172.

Liu, D., Ahmet, A., Ward, L., Krishnamoorthy, P., Mandelcorn, E. D., Leigh, R., Brown, J. P., Cohen, A. & Kim, H. 2013. A practical guide to the monitoring and management of the complications of systemic corticosteroid therapy. Allergy Asthma Clin Immunol, 9, 30.

Miguel-Alvarez, M., Santos-Lozano, A., Sanchis-Gomar, F., Fiuza-Luces, C., Pareja-Galeano, H., Garatachea, N. & Lucia, A. 2015. Non-steroidal anti-inflammatory drugs as a treatment for Alzheimer’s disease: a systematic review and meta-analysis of treatment effect. Drugs Aging, 32, 139–47.

Owais Qureshi, A. D. 2023. COX Inhibitors. StatPearls [Internet]: StatPearls Publishing.

Palumbo, M. L., Moroni, A. D., Quiroga, S., Castro, M. M., Burgueno, A. L. & Genaro, A. M. 2021. Immunomodulation induced by central nervous system-related peptides as a therapeutic strategy for neurodegenerative disorders. Pharmacol Res Perspect, 9, e00795.

Schwartz, D. M., Kanno, Y., Villarino, A., Ward, M., Gadina, M. & O’shea, J. J. 2017. JAK inhibition as a therapeutic strategy for immune and inflammatory diseases. Nat Rev Drug Discov, 16, 843–862.

Sharman, A. H. T. 2023. Corticosteroids. Treasure Island (FL): StatPearls.

Smolen, J. S., Aletaha, D. & Mcinnes, I. B. 2016. Rheumatoid arthritis. Lancet, 388, 2023–2038.

Stephenson, J., Nutma, E., Van Der Valk, P. & Amor, S. 2018. Inflammation in CNS neurodegenerative diseases. Immunology, 154, 204–219.

Strzelec, M., Detka, J., Mieszczak, P., Sobocinska, M. K. & Majka, M. 2023. Immunomodulation-a general review of the current state-of-the-art and new therapeutic strategies for targeting the immune system. Front Immunol, 14, 1127704.

Torrandell-Haro, G., Branigan, G. L., Vitali, F., Geifman, N., Zissimopoulos, J. M. & Brinton, R. D. 2020. Statin therapy and risk of Alzheimer’s and age-related neurodegenerative diseases. Alzheimers Dement (N Y), 6, e12108.

U.S. Food and Drug Administration (FDA). 2020. What are “biologics” questions and answers. [Online]. Available: https://www.fda.gov/about-fda/center-biologics-evaluation-and-research-cber/what-are-biologics-questions-and-answers [Accessed].

Varatharaj, A. & Galea, I. 2017. The blood-brain barrier in systemic inflammation. Brain Behav Immun, 60, 1–12.

Walsh, G. 2018. Biopharmaceutical benchmarks 2018. Nat Biotechnol, 36, 1136–1145.

Wang, J., Tan, L., Wang, H. F., Tan, C. C., Meng, X. F., Wang, C., Tang, S. W. & Yu, J. T. 2015. Anti-inflammatory drugs and risk of Alzheimer’s disease: an updated systematic review and meta-analysis. J Alzheimers Dis, 44, 385–96.

Wieczorek, I. & Strosznajder, R. P. 2024. S1P receptor modulators affect the toxicity of amyloid beta oligomers in microglial and neuronal cells. Folia Neuropathol, 62, 348–361.

Zhang, W., Xiao, D., Mao, Q. & Xia, H. 2023. Role of neuroinflammation in neurodegeneration development. Signal Transduct Target Ther, 8, 267.

Zheng, J., Chen, D., Xu, J., Ding, X., Wu, Y., Shen, H. C. & Tan, X. 2021. Small molecule approaches to treat autoimmune and inflammatory diseases (Part III): Targeting cytokines and cytokine receptor complexes. Bioorg Med Chem Lett, 48, 128229.

